# Assessing the Quality of a Personalized Prompt Generator and AI-Chatbot (ChatGPT) for Dietary and Exercise Planning in Obese Adults Using the Fuzzy Delphi Method

**DOI:** 10.1101/2025.08.27.25334528

**Authors:** Azwa Suraya Mohd Dan, Adam Linoby, Sazzli Shahlan Kasim, Sufyan Zaki, Razif Sazali, Yusandra Yusoff, Zulqarnain Nasir, Amrun Haziq Abidin

**Affiliations:** Faculty of Sports Science and Recreation, Universiti Teknologi MARA, Seremban Campus, Negeri Sembilan Branch, Negeri Sembilan, MALAYSIA; Faculty of Medicine, Universiti Teknologi MARA, Sungai Buloh Campus, Selangor Branch, Selangor, MALAYSIA; Faculty of Sports Science and Recreation, Universiti Teknologi MARA, Shah Alam Campus, Selangor Branch, Selangor, MALAYSIA

**Keywords:** Obesity Management, Artificial Intelligence, Exercise, Nutrition, Delphi Technique

## Abstract

**Background:** The potential of artificial intelligence (AI) to personalize dietary and exercise advice for obesity management is increasingly evident. However, the effectiveness and appropriateness of AI-generated recommendations hinge significantly on input quality and structured guidance. Despite growing interest, there remains a notable gap regarding a robust and validated prompt-generation mechanism designed explicitly for obesity-related lifestyle planning.

**Objectives:** This study aimed to evaluate and refine the quality of a personalized AI-driven framework (NExGEN-ChatGPT) for dietary and exercise prescriptions in obese adults, employing the Fuzzy Delphi Method (FDM) to capture and integrate expert consensus.

**Methods:** A multidisciplinary expert panel, comprising 21 professionals from nutrition, medicine, psychology, fitness, and AI domains, was engaged in this study. Using structured questionnaires, the experts systematically assessed and refined six primary constructs, further detailed into several evaluative elements, resulting in the consensus validation of 111 specific criteria.

**Results:** Findings identified critical consensus-driven standards essential for personalized, safe, and feasible obesity management through AI. Moreover, the study revealed prioritized criteria pivotal for maintaining practical relevance, safety, and high-quality personalized recommendations.

**Conclusions:** Consequently, this validated framework provides a substantial foundation for subsequent real-world application and further research, thereby enhancing the effectiveness, scalability, and individualization of obesity interventions leveraging AI.

## 1. Introduction

Globally, obesity has emerged as a profound public health crisis, with its prevalence steadily escalating over the last several decades. Recent estimates indicate that more than 650 million adults are obese worldwide, marking a near-tripling since 1975.^1^ Notably, adult obesity prevalence has exceeded 40% in countries like the United States, substantially increasing healthcare expenditures and economic burdens due to obesity-related conditions. It is projected that obesity-driven healthcare costs and productivity losses could surpass 3% of global GDP by 2030.^2^ Traditional one-to-one dietary and exercise counseling, although clinically effective (resulted in 5–10% body weight reduction over six to twelve months) remains resource-intensive and economically burdensome.^3^ Consequently, there is a compelling need for scalable alternatives capable of delivering personalized guidance to vast populations. Artificial Intelligence (AI)-driven conversational agents, such as ChatGPT, have recently attracted attention as viable solutions, offering personalized nutrition and physical activity coaching accessible 24/7, transcending human counselor availability constraints.^4^ Behavior-change theories, including the Capability, Opportunity, Motivation-Behavior (COM-B) model and Social Cognitive Theory, underpin the development of these chatbots, enhancing their capacity to facilitate effective, scalable lifestyle interventions.^5^

Empirical investigations assessing AI chatbot-generated dietary and exercise recommendations demonstrate promising alignment with established clinical guidelines. Recent comparative analyses suggest chatbot-generated weekly exercise, and caloric targets align closely with recommendations from authoritative bodies such as the World Health Organization and the American College of Sports Medicine.^6^ These AI-generated dietary plans typically adhere to recommended macronutrient balances, indicating basic safety and appropriateness. User engagement studies further underscore the potential of chatbots in weight management, highlighting sustained interactions driven by their accessibility and personalized feedback capabilities, resulting in weight-loss outcomes comparable to traditional counseling over short durations.^7,8^ Nevertheless, significant concerns persist regarding the accuracy and safety of chatbot-generated advice. Documented incidents reveal cases where health chatbots inadvertently provided unsafe dietary recommendations or misinformation, necessitating rigorous validation, clear safety protocols, and oversight mechanisms prior to extensive deployment.^9,10^

Systematic reviews and meta-analyses examining conversational AI agents for lifestyle modification reveal modest yet positive impacts on diet, physical activity, and body weight among obese populations. Recent meta-analyses have shown users of health chatbots typically achieve approximately 2–3 kg greater weight loss over three months compared to minimal interventions.^11^ Despite encouraging findings, considerable variability in study methodologies and limited long-term evaluations (typically under six months) present substantial knowledge gaps concerning the durability of behavior changes facilitated by AI interventions.^12,13^ Furthermore, the refinement of chatbot responses through advanced prompt-engineering strategies has shown potential to enhance personalization and clinical alignment of AI-generated advice, yet structured methods for optimizing these interactions remain nascent and insufficiently validated within obesity management.^14,15^

To systematically address these critical quality concerns in AI-generated dietary and exercise advice, expert consensus methods such as the Fuzzy Delphi Method (FDM) are increasingly employed. This method incorporates fuzzy logic to quantify expert agreement on essential criteria for health technology evaluations, effectively managing uncertainty in subjective judgments.^16^ Prior applications of FDM in telehealth and mobile health app evaluations have successfully established robust, expert-derived quality frameworks.^17,18^ Recently developed instruments, including the Quality Assessment of Medical AI (QAMAI) scale, highlight efforts toward standardized evaluations of medical AI; however, domain-specific criteria for nutrition and fitness recommendations are still lacking, particularly regarding personalization, nutritional adequacy, and safety tailored explicitly for obesity management.^19^

The Nutrition and Exercise Planning Artificial Intelligence Prompt Generator (NExGEN) is an online platform specifically developed to generate personalized dietary and exercise planning prompts for integration with AI-driven chatbots. NExGEN is designed to support individualized health and weight management goals by systematically incorporates diverse user-specific factors such as nutritional needs, fitness levels, anthropometric data, medical history, socioeconomic status, geographic location, stress levels, sleep patterns, and motivational aspects into its framework. Utilizing both offline (Microsoft Excel) and online spreadsheet applications (Microsoft Excel Online and Google Sheets), NExGEN produces highly tailored prompts designed explicitly for large language models such as ChatGPT, thereby facilitating precise AI-generated recommendations. The integration between NExGEN and ChatGPT capitalizes on AI’s decision-making capabilities to deliver personalized nutrition and exercise plans, promoting user autonomy and sustained adherence to lifestyle changes. Figure 1 illustrates the architecture framework of the NExGEN–ChatGPT system integration.

**Figure 1:** Architecture framework of NExGEN–ChatGPT system integration.

This research aims to rigorously assess and refine a novel integrated framework combining the NExGEN with ChatGPT, employing the FDM to derive expert consensus on the quality of generated lifestyle plans. Through iterative refinement, this approach seeks to validate the practicality, personalization, and accuracy of AI-driven dietary and exercise recommendations, providing a scalable, expert-validated solution to enhance obesity management outcomes among adults.

## 2. Methods

This study adopted a structured quantitative design, employing the FDM to systematically refine and validate the NExGEN-ChatGPT prompt generation framework for delivering personalized fitness and nutritional recommendations. The FDM was specifically selected due to its unique combination of iterative consensus-building typical of traditional Delphi,^20^ enhanced by fuzzy set theory, which accommodates subjective expert opinions and uncertainties inherent in interdisciplinary fields such as nutrition, fitness, psychology, medicine, and AI. In this approach, experts’ qualitative judgments, initially captured using Likert scales, were translated into quantitative Triangular Fuzzy Numbers represented by minimum (m1), moderate (m2), and maximum (m3) values. This allowed for capturing the nuanced and varying levels of agreement among experts effectively, thereby ensuring the accuracy and robustness of consensus outcomes in refining the framework.

### 2.1 NExGEN-ChatGPT Design and Process Flow

The initial phase of the NExGEN-ChatGPT prompt generation process begins with the participant completing a comprehensive initial survey. This survey collects extensive and specific personal data across multiple relevant areas, including nutritional habits, fitness status, anthropometric measurements, demographics, socioeconomic factors, medical and physiological health history, geographical location, sleep patterns, stress levels, and motivational influences. Comprising 111 targeted questions structured across 12 distinct elements, this survey aims to establish an individualized baseline that informs the personalization of fitness and nutritional plans through ChatGPT. Crucially, participants first define specific personal health objectives, including desired outcomes such as targeted weight loss, timelines, and the starting dates of their weight management programs (Figure 2). Detailed demographic and physiological measurements (e.g., body fat percentage, lean body mass, and heart rate) are collected alongside tailored questions concerning medical history, sleep, and stress management practices, further ensuring that the resulting personalized plans comprehensively reflect the individual’s specific needs.

**Figure 2:** Starting interface of NExGEN initial survey.

Following survey completion, participants encounter the first critical step in the NExGEN system, designated as the ‘1st Prompt’. This phase integrates the personalized data captured from the initial survey with a pre-defined narrative context designed explicitly to optimize interactions with ChatGPT. Participants copy this combined information which consist of detailed personal attributes alongside structured instructions into ChatGPT (using o3-mini model; the highest reasoning model at the time of data collection), thus enabling the chatbot to function effectively as a personal trainer, nutritionist, and health advisor (Figure 3). This carefully structured process ensures the chatbot’s responses remain consistent and informed throughout subsequent interactions, establishing a foundation for continuous, data-driven engagement tailored specifically to the individual’s evolving health journey.

**Figure 3:** Example of the 1st prompt page in NExGEN (left) and transposed data in ChatGPT (right).

Subsequent prompts further build upon this initial setup to address various aspects of personalized health management. The 2nd Prompt specifically targets lifestyle and behavioral modifications, offering detailed recommendations on diet, physical activities, and psychological well-being. The 3rd Prompt generates a customized weekly exercise plan tailored to the individual’s profile and goals, complete with locations, timings, and specific exercise instructions. The 4th Prompt develops a detailed weekly dietary plan, structured around personalized nutrition needs, accessible food choices, and clearly defined caloric intakes. Subsequently, the 5th Prompt provides a categorized grocery shopping list designed to support adherence to the dietary plan, specifying necessary ingredients and feasible substitutions. Finally, the 6th Prompt delivers an easy-to-follow meal preparation guide, featuring user-friendly recipes, portion guidance, cooking tips, and food safety advice, thereby equipping users with practical tools to effectively manage their nutrition and maintain consistent progress toward their goals.

### 2.2 Research Instrument

The primary instrument utilized in this research was a comprehensive self-administered questionnaire developed specifically for the FDM evaluation process. This bilingual questionnaire, available in Malay and English, was meticulously structured to facilitate clear understanding and accurate responses from the expert panel. It encompassed six distinct constructs, consisting of 111 specific items categorized into twelve critical evaluative elements relevant to fitness and nutritional planning. These constructs included detailed questions addressing demographic characteristics, physiological measurements, dietary habits, physical activity levels, psychological well-being, and behavioral motivations. Reliability of the instrument was established through cognitive pilot,^21^ achieving Cronbach’s alpha coefficients ranging from 0.714 to 0.891, thereby confirming the questionnaire’s internal consistency and suitability for expert.^22^

### 2.3 Data Collection and Preliminary Processing

The process of data collection and analysis is based on the implementation steps of FDM, as shown in Figure 4. Ethical clearance was secured from the Universiti Teknologi MARA Institutional Review Board (REC/09/2024 (PG/FB/30)), and the study was prospectively registered with the UMIN Trials Registry (UMIN000053570). Each prospective expert was approached individually, provided with a study briefing, and asked to sign an informed-consent form before receiving the questionnaire. To accommodate varying schedules and continuing public-health restrictions, questionnaires were distributed either face-to-face in hard copy or electronically as password-protected PDFs sent via e-mail or WhatsApp. All respondents completed the survey independently without researcher guidance, ensuring that responses reflected authentic, uninfluenced judgments.

**Figure 4:** Schematic diagram of FDM threshold.

#### 2.3.1 Step 1: Selection of Experts

A purposive sampling strategy guided the selection of 21 multidisciplinary experts, each holding a minimum of ten years’ professional.^23^ The panel included seasoned nutritionists, exercise specialists, medical practitioners, clinical psychologists, and AI researchers affiliated with academia, public healthcare institutions, and private sectors across Malaysia. This selection approach ensured comprehensive expert coverage across relevant disciplines, providing diverse and highly specialized insights crucial for refining the NExGEN-ChatGPT framework. Table 1 shows the demographic information and experts involved in this study.

**Table 1:** Experts’ demography.

| **Expert Category** | **Position** | **Level of Education** | **Years of Experience** |
| --- | --- | --- | --- |
| Accredited Nutritionist | Chief Dietitian at a hospital in the Kuala Lumpur, Malaysia. | Master | 19 Years |
|  | Nutritionist at a wellness center in Selangor, Malaysia. | Doctorate | 12 Years |
|  | Head of Nutrition at a clinic in Penang, Malaysia. | Doctorate | 18 Years |
|  | Senior Nutritionist at a health center in Ipoh, Malaysia. | Master | 14 Years |
|  | Nutritionist at a healthcare facility in Petaling Jaya, Malaysia. | Doctorate | 12 Years |
| Exercise Specialist | Fitness Director at a commercial fitness center in Kuala Lumpur, Malaysia. | Master | 12 Years |
|  | Exercise Physiologist at Malaysia Sports Institute. | Bachelor | 11 Years |
|  | Sports Therapist at a sports center in Selangor, Malaysia. | Master | 10 Years |
|  | Wellness Coach at a fitness studio in Johor, Malaysia. | Master | 11 Years |
|  | Exercise and Rehabilitation Specialist at a rehabilitation clinic in Johor, Malaysia. | Doctorate | 13 Years |
| Medical Personnel | General Practitioner, at a public hospital in Serdang, Malaysia. | Doctorate | 18 Years |
|  | General Practitioner at a private hospital in Johor, Malaysia. | Doctorate | 15 Years |
|  | General Practitioner at a health center in Kuala Lumpur, Malaysia. | Doctorate | 12 Years |
|  | Cardiologist at a public hospital in Penang, Malaysia. | Doctorate | 11 Years |
|  | Cardiologist at a heart institute in Malacca, Malaysia. | Doctorate | 16 Years |
| Accredited Psychologist | Educational Psychologist at a university in Penang, Malaysia. | Master | 10 Years |
|  | Research Psychologist at a university in Johor, Malaysia. | Doctorate | 12 Years |
|  | Clinical Psychologist at a health clinic in Selangor, Malaysia. | Master | 11 Years |
|  | Organizational Psychologist in Selangor, Malaysia. | Doctorate | 10 Years |
| AI Researcher | AI Lab Director at a public university in Kuala Lumpur, Malaysia. | Doctorate | 10 Years |
|  | Machine-learning Specialist at an AI-based company in Penang, Malaysia. | Master | 12 Years |

The expert panel comprised of accredited nutritionists (n = 5), exercise specialists (n = 5), medical practitioners (n = 5), accredited psychologists (n = 4), and AI-researchers (n = 2). The panel was diverse in terms of gender, with 11 males and 10 females participating. The ages of the experts ranged from 36 to 58 years, with a mean age of 47.3 ± 7.6 years. This demographic data was systematically collected at the beginning of the study to ensure a broad range of insights and experiences, essential for the FDM applied in refining the AI-driven health management tools.

#### 2.3.2 Step 2: Questionnaire Development

The development of the questionnaire followed a rigorous, iterative, and evidence-informed approach. Initially, extensive literature reviews were conducted to map relevant theoretical constructs in personalized nutrition and exercise planning. Semi-structured expert interviews were subsequently performed to identify emerging factors not fully covered in existing literature. Potential constructs encompass user interaction with fitness and nutrition programs, tailored behavioral modification recommendations, and engagement’s psychological effects. In developing the questionnaire, these constructs are dissected into quantifiable variables for examination via targeted questions. This step considers the operationalization of each variable in a manner that aligns with the study’s goals and is clear to the participating experts. The synthesized findings were then meticulously operationalized into precise, measurable questionnaire items. The final instrument balanced quantitative Likert-scale assessments with qualitative open-ended questions to obtain both measurable ratings and in-depth insights necessary for robust Delphi analysis.

#### 2.3.3 Step 3: Data Collection

Experts completed the finalized questionnaire within a specified two-week timeframe, facilitated by scheduled follow-ups to ensure timely submissions. This structured approach allowed for comprehensive yet independent expert evaluations, minimizing biases from researcher interventions. Following the acquisition of consent, the questionnaire was provided in both hardcopy format for face-to-face meetings and in a softcopy PDF format for those unable to meet in person. Responses were systematically collected and securely documented, ensuring consistency and integrity throughout the data collection phase.

#### 2.3.4 Step 4: Data Analysis

##### a) Fuzzy Delphi Scale

Expert evaluations collected via the Likert scales were converted to Triangular Fuzzy Numbers using a seven-point fuzzy scale to enhance analytical granularity. This conversion process effectively quantified the subjective expert judgments, facilitating precise aggregation and subsequent.^24,25^ The number of levels on the Fuzzy scale is represented by odd numbers. A higher Fuzzy ranking indicates greater accuracy of the obtained data. Figure 5 illustrates the mean triangular graph plotted against the Triangular values, specifically the three components of the Triangular Fuzzy Number.

**Figure 5:** Flowchart of FDM implementation for NExGEN-ChatGPT framework criteria evaluation.

##### b) Consensus Evaluation

The data analysis focuses on the use of Triangular Fuzzy Numbers to determine the value of the Threshold (d), which is important for achieving a level of expert agreement, as emphasized by.^26^ In line with,^27^ The degree of expert agreement was rigorously assessed using the vertex method to calculate inter-expert distances, setting a stringent d of ≤ 0.20 for acceptance. The vertex method was utilized to calculate the distance between the statistic for reporting ranked responses averages, enabling precise measurement of consensus among the experts. The distances for each Fuzzy number m = (m1, m2, m3) and n = (n1, n2, n3) calculated using the formula:

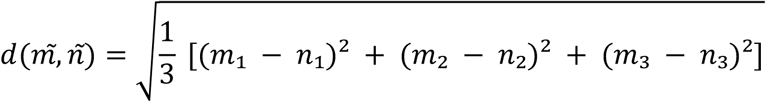

Criteria were further validated by requiring at least a 75% consensus rate among experts. This dual-threshold system ensured robust methodological rigor, significantly enhancing the validity of the consensus-driven outcomes.^27,28^

##### c) Defuzzification

The final analytical step involved defuzzification, employing the weighted average formula to convert aggregated fuzzy values into clear and actionable scores. Criteria achieving a defuzzified score (Amax; A) of 0.50 or higher were retained, reflecting adequate expert consensus and while those below the threshold were systematically excluded.^29^ The significance of the Fuzzy score (A) is important in establishing the ranking and priority of the elements under scrutiny. To calculate the Fuzzy score value (A), the formula Amax = 1/4 x (a1 + 2am + a2) was applied. The α-cut value, serving as a critical benchmark, is computed as the median of ‘0’ and ‘1’, resulting in α-cut = (0 + 1)/2 = 0.5. If the calculated Fuzzy score is found to be below this α-cut threshold, the criteria was deemed unacceptable, as this indicates a lack of expert agreement. This comprehensive defuzzification ensured the framework’s practical applicability and precision, readying the refined NExGEN-ChatGPT prompt generation system for subsequent validation studies and practical implementation.

## 3. Results

The findings are explained according to constructs, elements, and criteria compiled based on discussions with experts. Overall, all the criteria accepted for each construct and element met the requirements and conditions to pass the expert consensus of >75%, the value of Threshold (d) < 0.2 and α -cut is more than 0.5.

### 3.1 Construct 1: Goal Settings

Construct 1 focuses on goal setting and consists of three elements and six criteria. The elements specify the desired outcome for weight management, the targeted weight change, and the timeline for achieving these goals. Findings from the FDM analysis are reported in a table that lists only the criteria that met the acceptance threshold.

#### 3.1.1 Element: Desired Weight Management Outcomes

Table 2 highlights collective judgment on each criterion’s importance and applicability for ‘Desired Weight Management Outcomes’. The most significant criterion identified is the thorough initial and ongoing assessment of physical activity, which achieved unanimous expert agreement (100%), a threshold value of 0.127, and a high fuzzy score of 0.859. This underscores consistent assessment as essential for program effectiveness. Ranking second, utilizing specific metrics to measure improvements received a 90.48% consensus and a fuzzy score of 0.821, reinforcing the necessity for measurable outcomes. Aligning physical activity habits with overall health and weight management goals ranked third (90.5%, fuzzy score 0.765), emphasizing alignment with broader health objectives. Effective tracking systems and participant feedback ranked fourth (95.24%, 0.762), highlighting supportive infrastructure’s role. Technology tools such as apps and wearables received lower but notable acceptance (76.19%, 0.665). Lastly, diverse physical activities catering to individual interests showed moderate consensus (81%, 0.667), emphasizing personalized approaches.

**Table 2.** Accepted Criteria Position by Ranking for the Element of Desired Weight Management Outcomes.

|  | **Condition of Triangular Fuzzy Numbers** |  | **Condition of Defuzzification Process** |  |  |
| --- | --- | --- | --- | --- | --- |
| **Item** | **Threshold Value, d** | **Percentage of Experts Group Consensus (%)** | **Fuzzy Scores (A)** | **Position** | **Expert Consensus** |
| How thoroughly are initial and ongoing assessments of physical activity levels integrated into the program? | 0.127 | 100.0% | 0.859 | 1 | Accepted |
| How effectively are physical activity habits integrated with overall health | 0.16 | 90.50% | 0.765 | 3 | Accepted |
| and weight management goals? |  |  |  |  |  |
| Does the program include a wide range of physical activities to cater to different interests and capabilities? | 0.151 | 81.00% | 0.667 | 5 | Accepted |
| Are there effective systems in place for tracking physical activity and providing feedback to participants? | 0.136 | 95.24% | 0.762 | 4 | Accepted |
| How is technology (e.g., apps, wearable devices) used to | 0.174 | 76.19% | 0.665 | 6 | Accepted |
| support and<br>enhance<br>engagement in<br>physical<br>activities? |  |  |  |  |  |
| Are specific<br>metrics used to<br>measure<br>improvements<br>or changes in<br>physical activity<br>habits? | 0.179 | 90.48% | 0.821 | 2 | Accepted |

#### 3.1.2 Element: Target Weight Management Outcomes

Table 3 presents the prioritized criteria for ‘Target Weight Management Outcomes’ derived through the FDM, emphasizing their importance and feasibility based on expert consensus percentages and fuzzy scores. The highest-ranked criterion, clearly and precisely defining target weight change goals, achieved unanimous expert consensus (100%) and a fuzzy score of 0.806, highlighting clarity and specificity as essential for effective weight management. The alignment of weight change goals with established health standards, such as BMI and body fat percentage, ranked second, receiving 90.48% consensus and a fuzzy score of 0.786, underscoring the need for evidence-based objectives. Goals tailored realistically to individual health and fitness status ranked third (76.2%, 0.724), reinforcing the value of practicality. Personalization based on age, gender, and lifestyle ranked fourth (76.19%, 0.656), further supporting individualized goal-setting approaches. The safety of weight change goals ranked fifth (80.95%, 0.640), while realistic timelines, despite a high consensus (90.5%), had a lower fuzzy score (0.576), suggesting timeline practicality is less foundational compared to other criteria.

**Table 3:** Accepted Criteria Position by Ranking for the Element of Target Weight Management Outcomes.

|  | **Condition of Triangular**<br>**Fuzzy Numbers** |  | **Condition of**<br>**Defuzzification**<br>**Process** |  |  |
| --- | --- | --- | --- | --- | --- |
| **Item** | **Threshold**<br>**Value, d** | **Percentage of**<br>**Experts**<br>**Group**<br>**Consensus**<br>**(%)** | **Fuzzy Scores**<br>**(A)** | **Position** | **Expert**<br>**Consensus** |
| Are the target weight change goals clearly and precisely defined? | 0.152 | 100.0% | 0.806 | 1 | Accepted |
| Are the target weight change goals realistic and feasible given the user's current health and fitness status? | 0.187 | 76.20% | 0.724 | 3 | Accepted |
| Are the weight management outcomes realistic and achievable within the specified timelines? | 0.180 | 90.50% | 0.576 | 6 | Accepted |
| Do the target weight change goals align with recognized health and medical standards (e.g., BMI, body fat percentage)? | 0.196 | 90.48% | 0.786 | 2 | Accepted |
| Are the target weight change goals personalized based on individual | 0.188 | 76.19% | 0.656 | 4 | Accepted |
| characteristics<br>such as age,<br>gender, health<br>conditions, and<br>lifestyle? |  |  |  |  |  |
| Are the target<br>weight change<br>goals set in a<br>manner that<br>ensures the<br>health and<br>safety of the<br>individual (e.g.,<br>avoiding rapid<br>weight loss/gain<br>that could be<br>harmful)? | 0.180 | 80.95% | 0.64 | 5 | Accepted |

#### 3.1.3 Element: Timeline for Achieving Goals

Table 4 presents criteria ranked through the FDM concerning timelines for ‘Timeline for Achieving Goals’, based on expert consensus and fuzzy scores, emphasizing their relative importance and practical relevance. The top-ranked criterion, assessing if timelines are realistic and achievable considering individual fitness levels and lifestyles, attained an 85.7% consensus and the highest fuzzy score of 0.725, indicating the importance of aligning timelines with personal capabilities. The second-ranked criterion, structuring timelines to sustain motivation and engagement, received identical consensus (85.71%) but a lower fuzzy score (0.595), underscoring motivation’s role throughout the goal process. Flexibility in adjusting goals and timeframes ranked third (80.95%, 0.562), reinforcing the need for adaptable planning. Fourth, incorporating incremental milestones for monitoring progress and adjustments scored 85.7% consensus and a fuzzy score of 0.557, highlighting structured checkpoints. Personalization of timelines based on age, gender, health, and lifestyle was fifth (85.71%, 0.530). Lastly, prioritizing health and safety by avoiding rapid changes ranked sixth (81%, 0.529), stressing cautious timeline planning.

**Table 4:** Accepted Criteria Position by Ranking for the Element of Timeline for Achieving Goals.

|  | **Condition of Triangular Fuzzy Numbers** |  | **Condition of Defuzzification Process** |  |  |
| --- | --- | --- | --- | --- | --- |
| **Item** | **Threshold Value, d** | **Percentage of Experts Group Consensus (%)** | **Fuzzy Scores (A)** | **Position** | **Expert Consensus** |
| Is the proposed timeline realistic and achievable based on the user's current fitness level and lifestyle? | 0.165 | 85.7% | 0.725 | 1 | Accepted |
| Does the timeline ensure that health and safety are prioritized, avoiding overly | 0.175 | 81.0% | 0.529 | 6 | Accepted |
| rapid or extreme changes? |  |  |  |  |  |
| Are there clear incremental milestones within the timeline to monitor progress and make adjustments? | 0.187 | 85.7% | 0.557 | 4 | Accepted |
| Does the timeline allow for flexibility to adjust goals and timeframes based on progress and unforeseen challenges? | 0.171 | 80.95% | 0.562 | 3 | Accepted |
| Is the timeline structured in a way that | 0.198 | 85.71% | 0.595 | 2 | Accepted |
| maintains<br>motivation and<br>engagement<br>over the<br>duration? |  |  |  |  |  |
| Is the timeline<br>personalized<br>according to<br>individual<br>factors such as<br>age, gender,<br>health status,<br>and lifestyle? | 0.174 | 85.71% | 0.530 | 5 | Accepted |

### 3.2 Construct 2: Demographics and Physical Characteristics

Construct 2 addresses demographic and physical characteristics. It comprises three elements: age; height and weight; and physiological indicators, specifically body-fat percentage and lean body mass.

#### 3.2.1 Element: Age

The results in Table 5 rank criteria concerning ‘Age’ in nutrition and exercise recommendations using the FDM. The top criterion, assessing whether strategies are safe across age groups given health risks, had the highest expert consensus at 95.24% and a fuzzy score of 0.681, emphasizing safety and risk mitigation. The second-ranked criterion addressed the clarity and appropriateness of recommendations for diverse age groups (95.2% consensus, 0.673 fuzzy score), highlighting the necessity of clear, adaptable guidelines. Adaptability to individual differences ranked third (80.95% consensus, 0.648 fuzzy score), underscoring personalized recommendations. Specific nutritional and exercise requirements for each age group ranked fourth (90.5% consensus, 0.603 fuzzy score). Consideration of developmental and physiological variations was fifth (85.7%, 0.576 fuzzy score), while meeting specific nutritional needs was sixth (85.71%, 0.511 fuzzy score), establishing foundational importance.

**Table 5.**
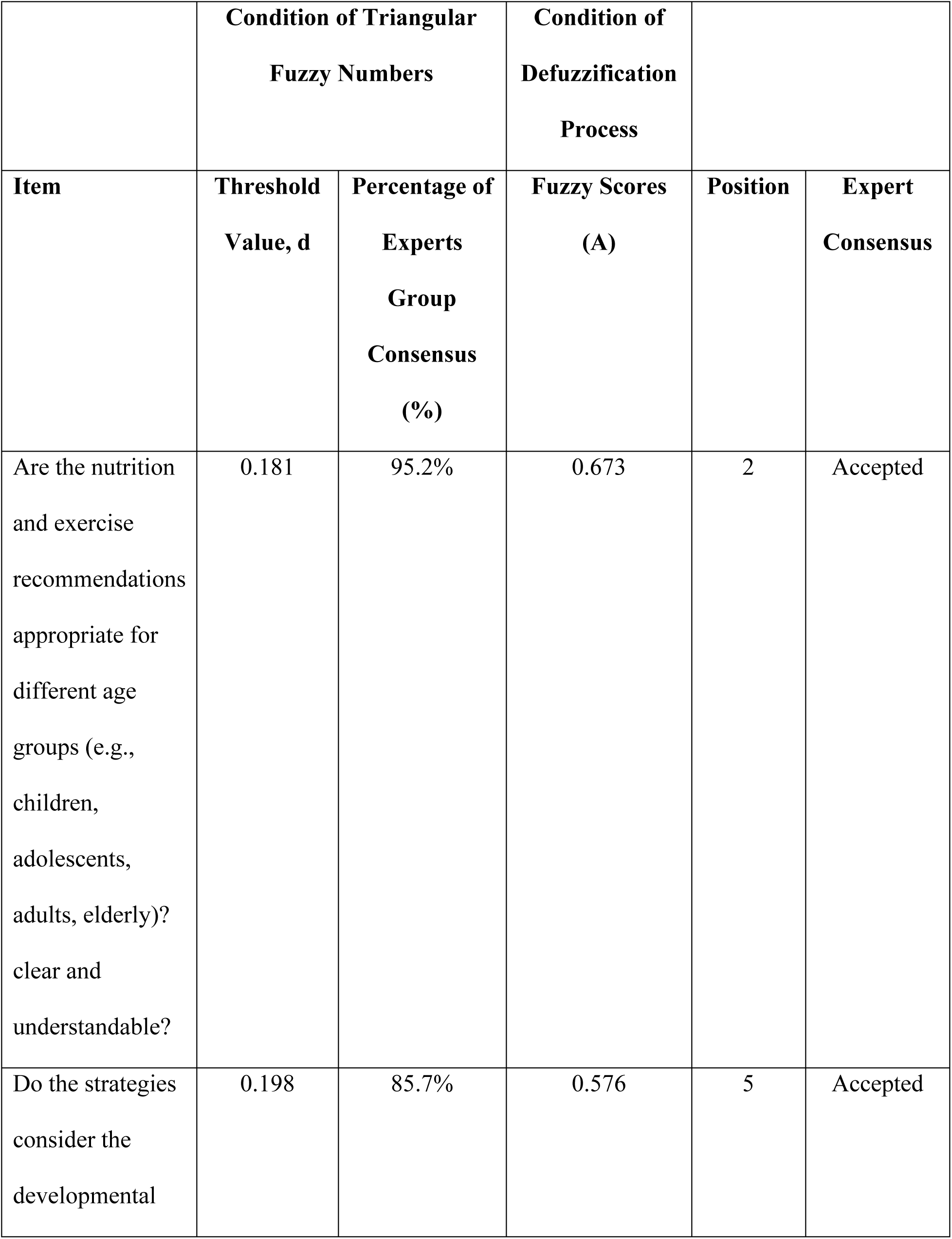
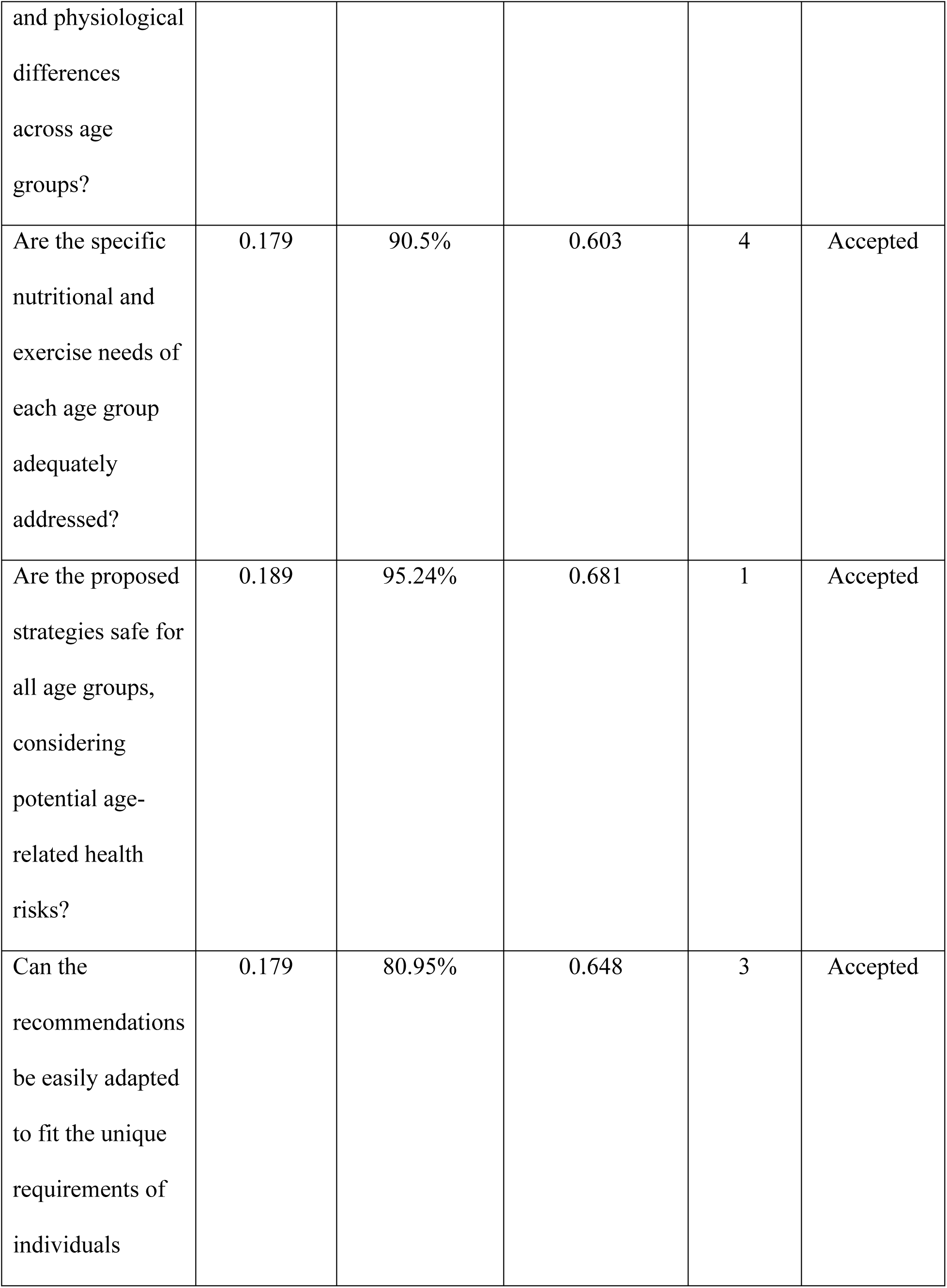
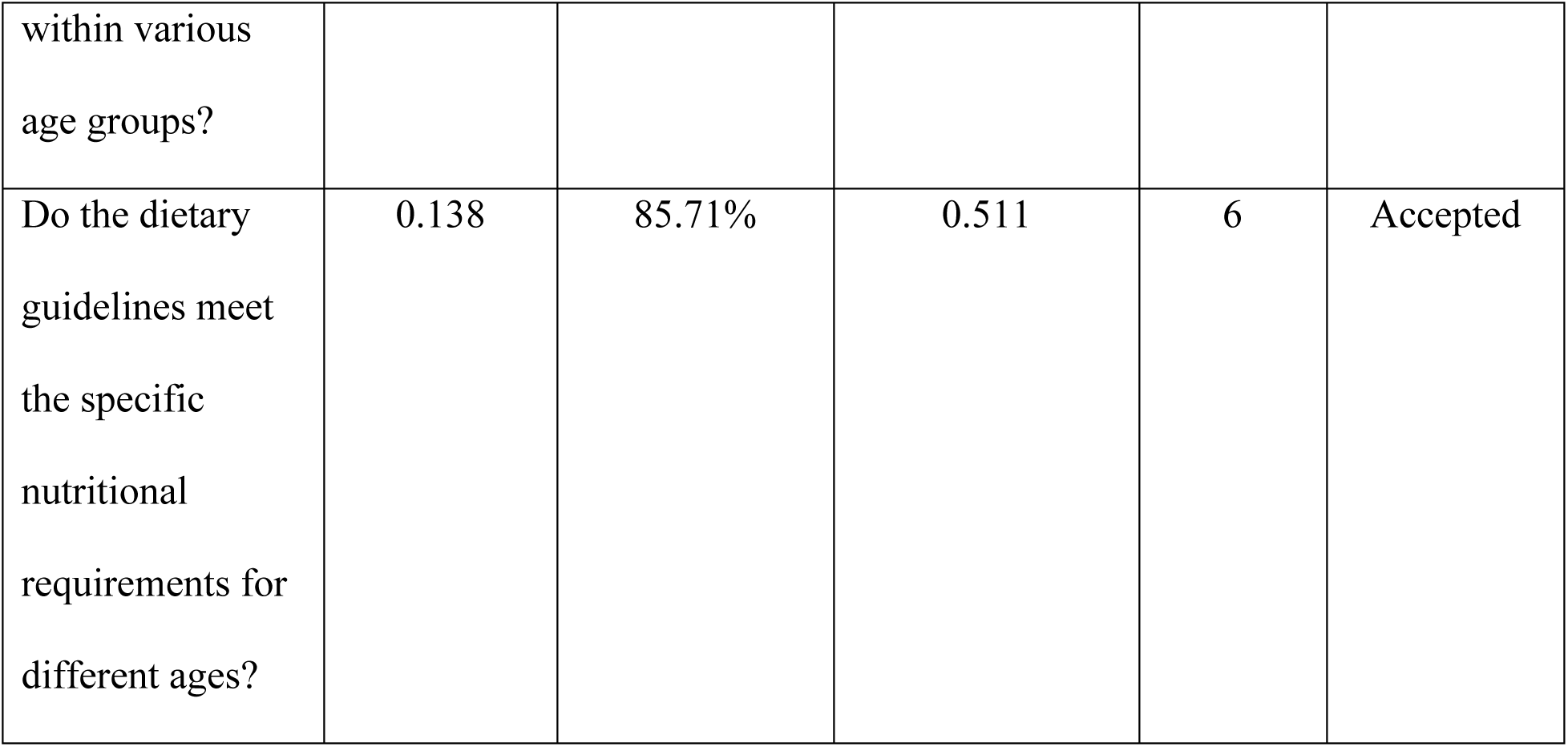
Accepted Criteria Position by Ranking for the Element of Age.

| **Item** | **Condition of Triangular Fuzzy Numbers** |  | **Condition of Defuzzification Process** |  |  |
| --- | --- | --- | --- | --- | --- |
|  | **Threshold Value, d** | **Percentage of Experts Group Consensus (%)** | **Fuzzy Scores (A)** | **Position** | **Expert Consensus** |
| Are the nutrition and exercise recommendations appropriate for different age groups (e.g., children, adolescents, adults, elderly)? clear and understandable? | 0.181 | 95.2% | 0.673 | 2 | Accepted |
| Do the strategies consider the developmental | 0.198 | 85.7% | 0.576 | 5 | Accepted |
| and physiological differences across age groups? |  |  |  |  |  |
| Are the specific nutritional and exercise needs of each age group adequately addressed? | 0.179 | 90.5% | 0.603 | 4 | Accepted |
| Are the proposed strategies safe for all age groups, considering potential age-related health risks? | 0.189 | 95.24% | 0.681 | 1 | Accepted |
| Can the recommendations be easily adapted to fit the unique requirements of individuals | 0.179 | 80.95% | 0.648 | 3 | Accepted |
| within various age groups? |  |  |  |  |  |
| Do the dietary guidelines meet the specific nutritional requirements for different ages? | 0.138 | 85.71% | 0.511 | 6 | Accepted |

#### 3.2.2 Element: Body Composition

The results presented in Table 6 prioritize criteria for nutrition and exercise recommendations based on ‘Body Composition’ using the FDM. The top-ranked criterion, addressing whether recommendations align with specific Body Mass Index (BMI) categories, achieved unanimous expert consensus (100%) and the highest fuzzy score (0.681), highlighting the importance of BMI-targeted strategies. The second criterion, assessing the safety of strategies across different height and weight profiles, also received full consensus (100%) and a fuzzy score of 0.605, emphasizing safety considerations. Personalization according to individual height and weight ranked third (95.2%, fuzzy score 0.576), underscoring tailored approaches. Realistic and achievable weight management goals placed fourth (85.71%, fuzzy score 0.557), ensuring practicality. Addressing nutritional needs across body sizes was fifth (90.48%, fuzzy score 0.517), emphasizing dietary inclusivity. Lastly, suitability of exercise intensity and type ranked sixth (76.19%, fuzzy score 0.511), indicating lower relative emphasis.

**Table 6.** Accepted Criteria Position by Ranking for the Element of Body Composition.

|  | **Condition of Triangular**<br>**Fuzzy Numbers** |  | **Condition of**<br>**Defuzzification**<br>**Process** |  |  |
| --- | --- | --- | --- | --- | --- |
| **Item** | **Threshold**<br>**Value, d** | **Percentage of**<br>**Experts**<br>**Group**<br>**Consensus**<br>**(%)** | **Fuzzy Scores**<br>**(A)** | **Position** | **Expert**<br>**Consensus** |
| Are the nutrition and exercise recommendations tailored to different Body Mass Index (BMI) categories (e.g., underweight, normal weight, overweight, obese)? | 0.186 | 100.0% | 0.681 | 1 | Accepted |
| Are the recommendations | 0.162 | 95.2% | 0.576 | 3 | Accepted |
| personalized<br>based on<br>individual height<br>and weight<br>measurements? |  |  |  |  |  |
| Are the proposed<br>strategies safe for<br>individuals with<br>different height<br>and weight<br>profiles,<br>considering<br>potential health<br>risks? | 0.152 | 100.0% | 0.605 | 2 | Accepted |
| Are the specific<br>nutritional needs<br>of individuals<br>with different<br>body sizes<br>adequately<br>addressed? | 0.164 | 90.48% | 0.517 | 5 | Accepted |
| Is the intensity and type of exercise recommended appropriate for different height and weight categories? | 0.193 | 76.19% | 0.511 | 6 | Accepted |
| Are weight management goals realistic and achievable for individuals with different height and weight profiles? | 0.187 | 85.71% | 0.557 | 4 | Accepted |

#### 3.2.3 Element: Physiological Measurements

Table 7 presents prioritized element related to ‘Physiological Measurements’. The highest-ranked criterion is evaluating whether physiological measurements ensure participant safety and health monitoring during nutrition and exercise programs. This criterion achieved an expert consensus of 85.7% and the top fuzzy score (0.659), highlighting participant safety. Ranking second, the criterion on personalized strategies based on individual physiological data also received 85.7% consensus (0.630 fuzzy score), emphasizing tailored interventions. Employing modern technologies to enhance physiological data collection, analysis, and feedback ranked third (90.48% consensus, 0.613 fuzzy score), underscoring technology’s role in data accuracy. Ensuring reliable and accurate measurement tools ranked fourth (95.2% consensus, fuzzy score 0.576). Adjusting fitness and nutrition plans according to changing physiological data was fifth (80.95%, 0.573), with tracking and reporting systems ranking sixth (80.95%, 0.511), reflecting their supporting roles.

**Table 7.**
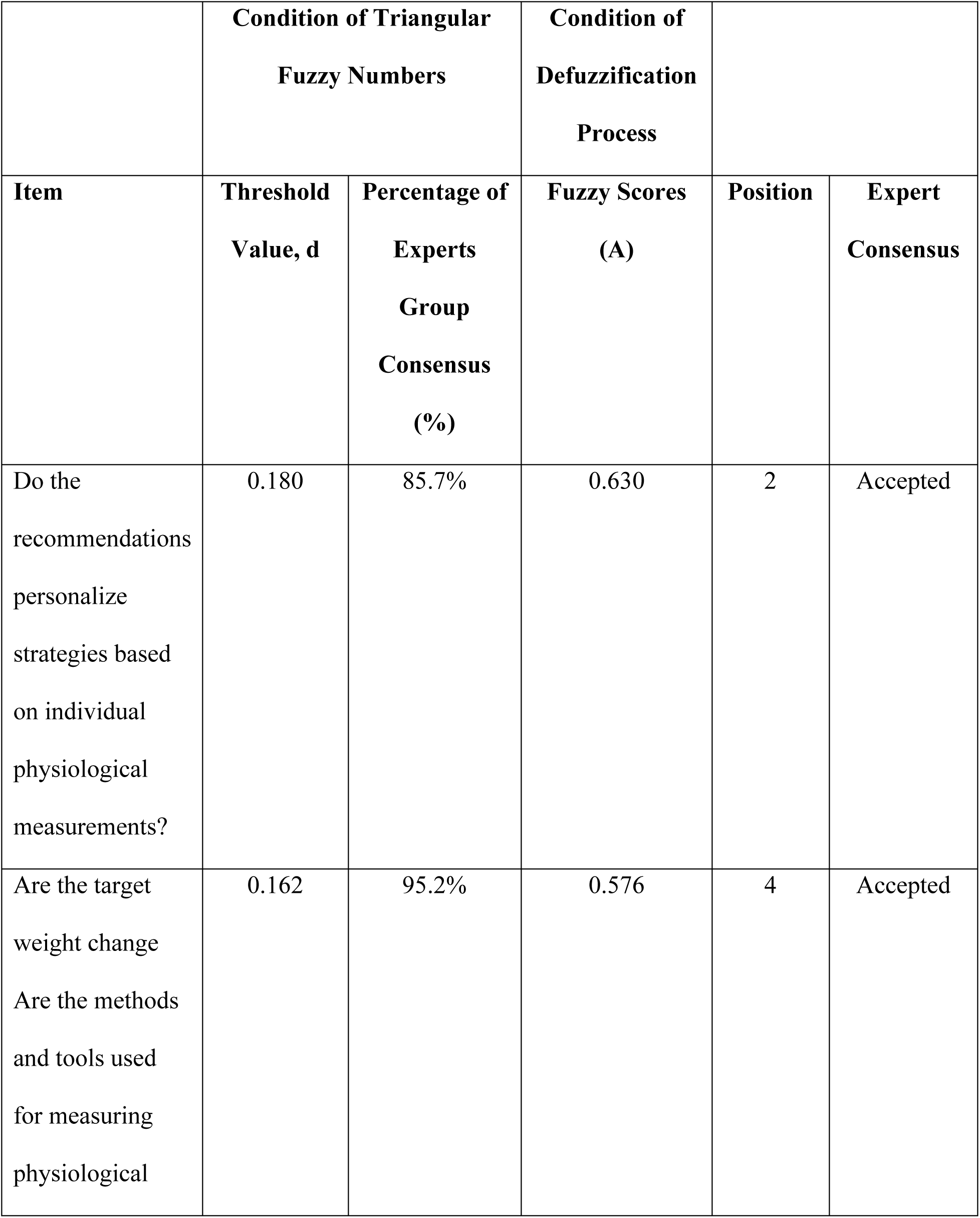
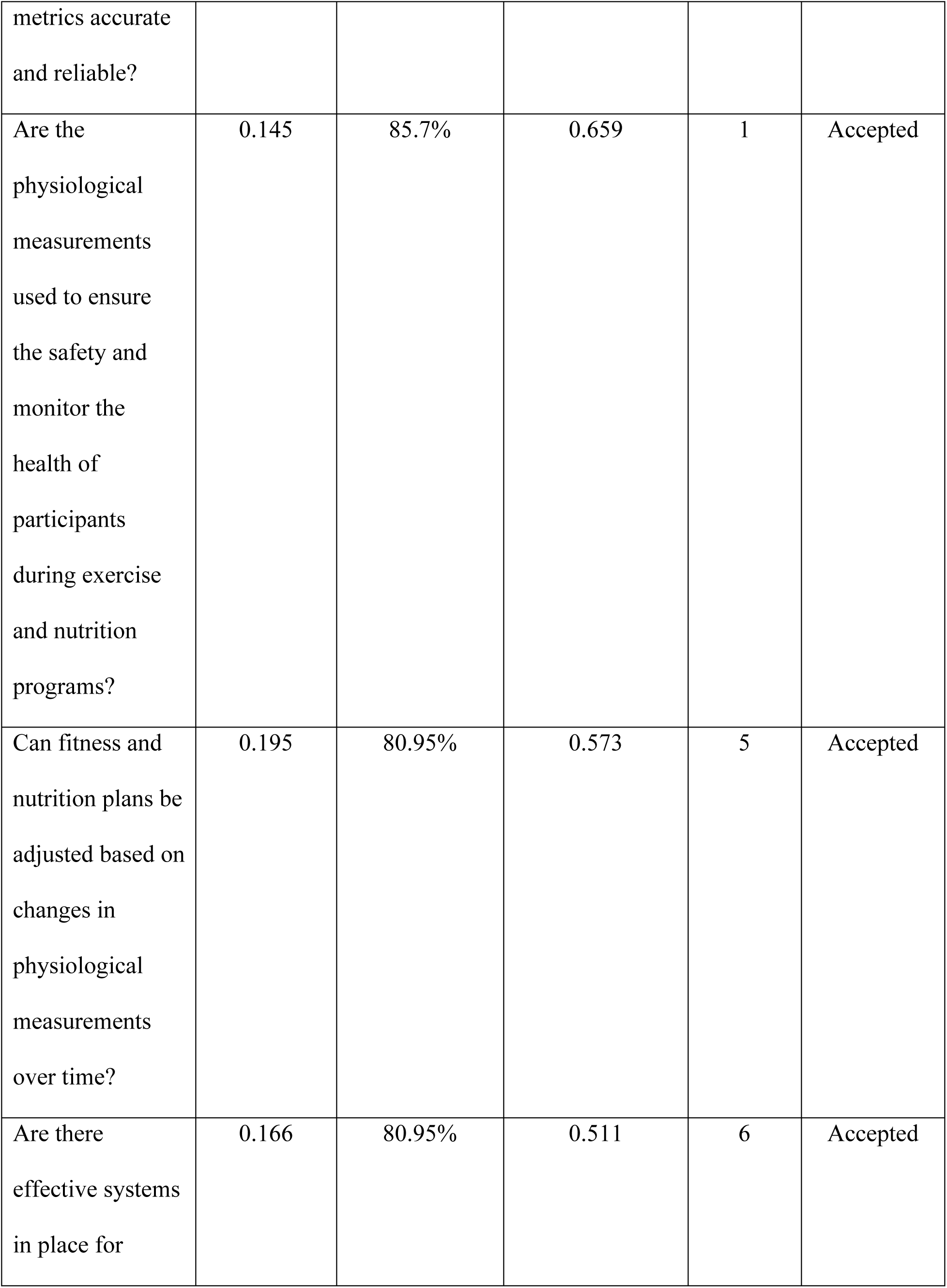
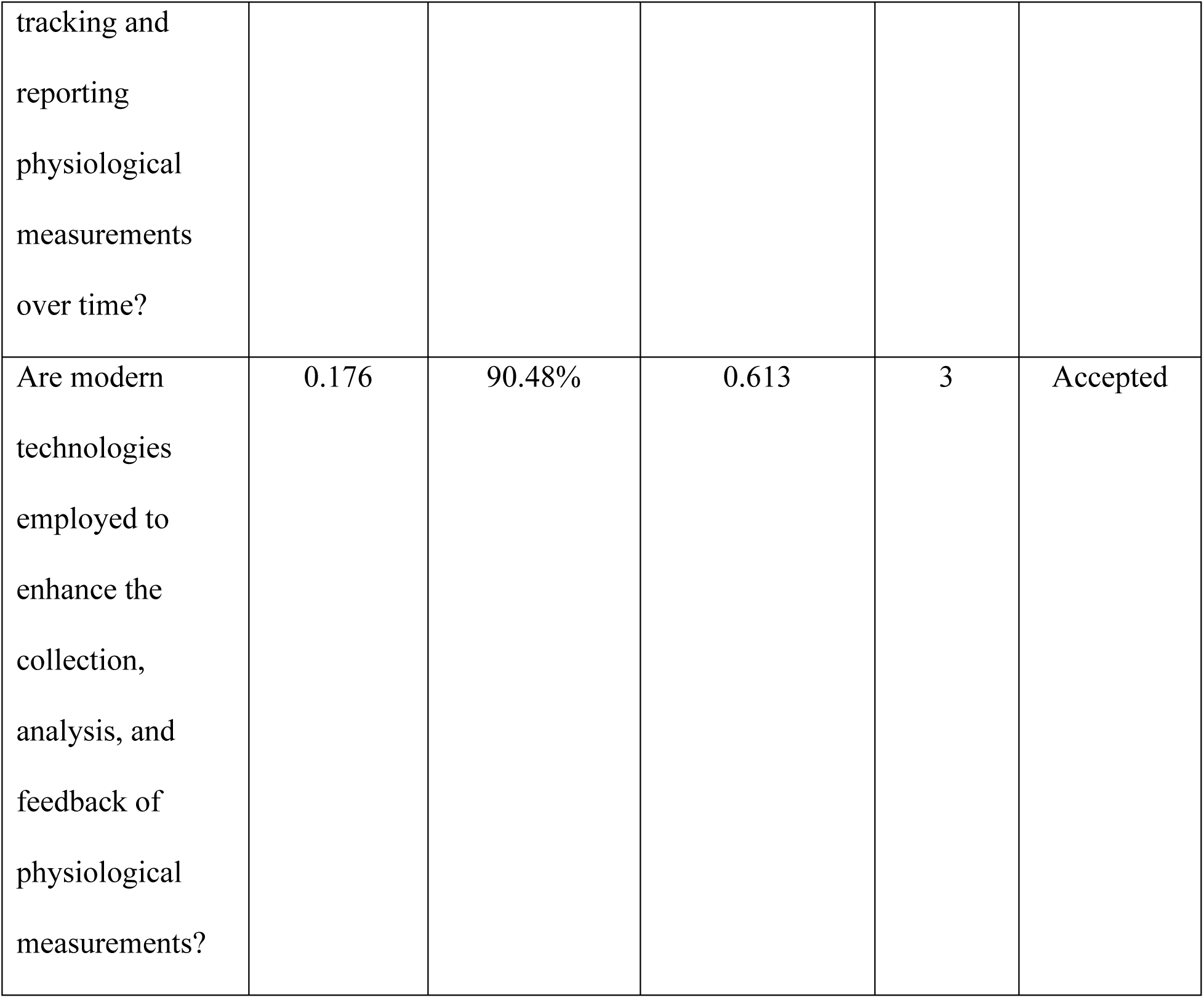
Accepted Criteria Position by Ranking for the Element of Physiological Measurements.

|  | **Condition of Triangular Fuzzy Numbers** |  | **Condition of Defuzzification Process** |  |  |
| --- | --- | --- | --- | --- | --- |
| **Item** | **Threshold Value, d** | **Percentage of Experts Group Consensus (%)** | **Fuzzy Scores (A)** | **Position** | **Expert Consensus** |
| Do the recommendations personalize strategies based on individual physiological measurements? | 0.180 | 85.7% | 0.630 | 2 | Accepted |
| Are the target weight change<br>Are the methods and tools used for measuring physiological | 0.162 | 95.2% | 0.576 | 4 | Accepted |
| metrics accurate and reliable? |  |  |  |  |  |
| Are the physiological measurements used to ensure the safety and monitor the health of participants during exercise and nutrition programs? | 0.145 | 85.7% | 0.659 | 1 | Accepted |
| Can fitness and nutrition plans be adjusted based on changes in physiological measurements over time? | 0.195 | 80.95% | 0.573 | 5 | Accepted |
| Are there effective systems in place for | 0.166 | 80.95% | 0.511 | 6 | Accepted |
| tracking and reporting physiological measurements over time? |  |  |  |  |  |
| Are modern technologies employed to enhance the collection, analysis, and feedback of physiological measurements? | 0.176 | 90.48% | 0.613 | 3 | Accepted |

### 3.3 Construct 3: Health and Medical History

The ‘Health and Medical History’ is the third construct with 2 element and 6 criteria for each element.

#### 3.3.1 Element: Medical Conditions

Table 8 presents analysis results related to ‘Medical Conditions,’ emphasizing tailored and inclusive strategies for fitness and nutrition management. The criterion advocating comprehensive inclusion of diverse medical conditions received 81% expert consensus and the highest fuzzy score (0.741), emphasizing broad applicability. Ranked second, customizing exercise and diet plans for specific medical conditions obtained unanimous consensus (100%, fuzzy score 0.683), highlighting personalized adaptation. Accessibility and feasibility for individuals with medical limitations ranked third (85.71%, fuzzy score 0.630), reinforcing inclusivity. Fourth-ranked was the presence of feedback mechanisms to assess program effectiveness in managing medical conditions (90.48%, fuzzy score 0.613), emphasizing continuous improvement. Safety considerations for participants with varying medical conditions ranked fifth (85.7%, fuzzy score 0.594), underlining risk reduction. Lastly, monitoring the impact of fitness and nutrition programs on medical conditions ranked sixth (80.95%, fuzzy score 0.530), underscoring adaptability based on changing health conditions.

**Table 8.** Accepted Criteria Position by Ranking for the Element of Medical Conditions.

|  | **Condition of Triangular**<br>**Fuzzy Numbers** |  | **Condition of**<br>**Defuzzification**<br>**Process** |  |  |
| --- | --- | --- | --- | --- | --- |
| **Item** | **Threshold**<br>**Value, d** | **Percentage of**<br>**Experts**<br>**Group**<br>**Consensus**<br>**(%)** | **Fuzzy Scores**<br>**(A)** | **Position** | **Expert**<br>**Consensus** |
| Are a wide range of known medical conditions considered in the fitness and nutrition recommendations (e.g., diabetes, heart disease, arthritis)? | 0.181 | 81.0% | 0.741 | 1 | Accepted |
| How effectively are the exercise and diet plans | 0.161 | 100.0% | 0.683 | 2 | Accepted |
| customized to accommodate specific medical conditions? |  |  |  |  |  |
| Are the recommendations safe for individuals with different medical conditions, minimizing health risks? | 0.194 | 85.7% | 0.594 | 5 | Accepted |
| Are there mechanisms in place to monitor the impact of the fitness and diet plans on the medical conditions and adjust them as necessary? | 0.199 | 80.95% | 0.530 | 6 | Accepted |
| Are the programs accessible and feasible for individuals with varying degrees of medical limitations? | 0.180 | 85.71% | 0.630 | 3 | Accepted |
| Is there a feedback system to gather input from participants on the effectiveness of the programs in managing their medical conditions? | 0.176 | 90.48% | 0.613 | 4 | Accepted |

#### 3.2.3 Element: Medications

Table 9 presents prioritized criteria related to the ‘Medications’ construct, emphasizing medication considerations in fitness and nutrition planning. The highest-ranked criterion which is ensuring exercise and nutrition recommendations avoid negative interactions with prescribed medications, achieved 95.2% consensus and the highest fuzzy score (0.698), underscoring the significance of safety. Second-ranked was effective customization of health plans based on medication effects (81% consensus, fuzzy score 0.667), highlighting personalized approaches. Adaptability of plans to accommodate medication regimen changes ranked third (76.2%, fuzzy score 0.663), emphasizing program flexibility. Fourth-ranked was responsiveness regarding necessary medication adjustments due to fitness and nutrition changes (95.24% consensus, fuzzy score 0.644). Ethical and legal considerations for medication-related privacy ranked fifth (85.71%, fuzzy score 0.622), reinforcing participant trust. Lastly, integrating medical records and medication histories into planning ranked sixth (80.95%, fuzzy score 0.529), indicating foundational yet secondary importance. Results emphasize safety, personalization, adaptability, and ethical compliance in health interventions.

**Table 9.** Accepted Criteria Position by Ranking for the Element of Medications.

|  | **Condition of Triangular**<br>**Fuzzy Numbers** |  | **Condition of**<br>**Defuzzification**<br>**Process** |  |  |
| --- | --- | --- | --- | --- | --- |
| **Item** | **Threshold**<br>**Value, d** | **Percentage of**<br>**Experts**<br>**Group**<br>**Consensus**<br>**(%)** | **Fuzzy Scores**<br>**(A)** | **Position** | **Expert**<br>**Consensus** |
| Are the exercise and nutrition recommendations designed to avoid negative interactions with commonly prescribed medications? | 0.181 | 95.2% | 0.698 | 1 | Accepted |
| How effectively are health and fitness plans customized to accommodate the | 0.151 | 81.0% | 0.667 | 2 | Accepted |
| effects of current medications on individuals? |  |  |  |  |  |
| Can the fitness and nutrition plans be easily adjusted based on changes in medication regimes? | 0.190 | 76.2% | 0.663 | 3 | Accepted |
| Are medical records that include medication histories integrated into the fitness and nutrition planning process? | 0.175 | 80.95% | 0.529 | 6 | Accepted |
| Are considerations for medication | 0.140 | 95.24% | 0.644 | 4 | Accepted |
| adjustments included if fitness and nutritional changes necessitate such adjustments? |  |  |  |  |  |
| Are ethical and legal considerations regarding medication information and privacy maintained throughout the program? | 0.183 | 85.71% | 0.622 | 5 | Accepted |

### 3.4 Construct 4: Lifestyle and Behaviour

#### 3.4.1 Element: Sleep Pattern

Table 10 highlights the element of ‘Sleep Pattern’. The highest-ranked criterion, adjusting programs for participants with diagnosed sleep disorders or irregular sleep patterns, achieved 90.48% consensus and a fuzzy score of 0.673, indicating strong agreement on addressing individual sleep-related challenges. Addressing the impact of sleep quality and duration on fitness and health outcomes ranked second (76.2% consensus, 0.665), reflecting sleep’s role in recovery, weight control, and energy. Educating participants on sleep’s connection to health ranked third (80.95%, 0.638). Integrating sleep patterns into personalized plans ranked fourth (81%, 0.637), emphasizing tailored approaches. Customized recommendations based on individual sleep quality ranked fifth (85.7%, 0.602), followed by monitoring systems for tracking sleep (85.71%, 0.583), highlighting data-driven approaches to improving health outcomes.

**Table 10.** Accepted Criteria Position by Ranking for the Element of Sleep Pattern.

|  | **Condition of Triangular**<br>**Fuzzy Numbers** |  | **Condition of**<br>**Defuzzification**<br>**Process** |  |  |
| --- | --- | --- | --- | --- | --- |
| **Item** | **Threshold**<br>**Value, d** | **Percentage of**<br>**Experts**<br>**Group**<br>**Consensus**<br>**(%)** | **Fuzzy Scores**<br>**(A)** | **Position** | **Expert**<br>**Consensus** |
| How well are sleep patterns considered and integrated into personalized fitness and nutrition plans? | 0.198 | 81.0% | 0.637 | 4 | Accepted |
| Are the effects of sleep quality and duration on health and fitness outcomes sufficiently | 0.174 | 76.2% | 0.665 | 2 | Accepted |
| addressed in the program? |  |  |  |  |  |
| How effectively are recommendations customized based on individual sleep quality and patterns? | 0.191 | 85.7% | 0.602 | 5 | Accepted |
| Are participants educated about the importance of sleep and its impact on weight management and overall health? | 0.189 | 80.95% | 0.638 | 3 | Accepted |
| Are there effective systems in place for monitoring and tracking changes in sleep patterns as part of the | 0.184 | 85.71% | 0.583 | 6 | Accepted |
| fitness and<br>nutrition<br>program? |  |  |  |  |  |
| Are adjustments<br>made for<br>participants with<br>diagnosed sleep<br>disorders or<br>irregular sleep<br>patterns? | 0.186 | 90.48% | 0.673 | 1 | Accepted |

#### 3.4.2 Element: Stress Level

The findings from Table 11 shows the prioritized criteria for ‘Stress Level’. The top-ranked criterion, considering stress levels in relation to health outcomes, such as weight management, achieved the highest expert consensus (95.2%) and fuzzy score (0.784), highlighting stress as a significant factor affecting physical and mental health. Ranked second, conducting initial and ongoing stress assessments to tailor program recommendations obtained 81% consensus and a fuzzy score of 0.738, emphasizing personalized evaluation. Integrating stress management into fitness and nutrition programs ranked third (76.2%, 0.663). Ensuring the safety and effectiveness of stress management methods ranked fourth (80.95%, 0.621), followed by promoting relaxation techniques like meditation and yoga (80.95%, 0.538). Customizing stress management based on individual triggers ranked sixth (80.95%, 0.529), supporting a holistic, personalized approach to wellness.

**Table 11.** Accepted Criteria Position by Ranking for the Element of Stress Level.

|  | **Condition of Triangular**<br>**Fuzzy Numbers** |  | **Condition of**<br>**Defuzzification**<br>**Process** |  |  |
| --- | --- | --- | --- | --- | --- |
| **Item** | **Threshold**<br>**Value, d** | **Percentage**<br>**of Experts**<br>**Group**<br>**Consensus**<br>**(%)** | **Fuzzy Scores**<br>**(A)** | **Position** | **Expert**<br>**Consensus** |
| Are initial and ongoing assessments of stress levels included in the program to tailor recommendations? | 0.182 | 81.0% | 0.738 | 2 | Accepted |
| How are stress levels considered in relation to their impact on health outcomes, including weight management? | 0.161 | 95.2% | 0.784 | 1 | Accepted |
| How effectively<br>are stress<br>management<br>strategies<br>integrated into<br>fitness and<br>nutrition plans? | 0.190 | 76.2% | 0.663 | 3 | Accepted |
| Are stress<br>management<br>techniques<br>customized based<br>on individual<br>stress triggers and<br>responses? | 0.175 | 80.95% | 0.529 | 6 | Accepted |
| Are the<br>recommended<br>stress<br>management<br>techniques safe<br>and proven<br>effective? | 0.198 | 80.95% | 0.621 | 4 | Accepted |
| Are relaxation techniques (e.g., meditation, deep breathing, yoga) incorporated into the program? | 0.188 | 80.95% | 0.538 | 5 | Accepted |

#### 3.4.3 Element: Physical Activity Habits

The results from Table 12 prioritize criteria for integrating ‘Physical Activity Habits’. The highest-ranked criterion, which is effectively aligning physical activity habits with overall health and weight management goals have achieved the top fuzzy score (0.781) and expert consensus (85.7%), emphasizing alignment with broader health objectives. The use of technology, such as apps and wearable devices, ranked second (85.71%, 0.760), highlighting its role in enhancing participant engagement. Consistent initial and ongoing assessments of physical activity levels ranked third (95.2%, 0.689), underscoring the importance of regular monitoring. Specific metrics to measure physical activity improvements ranked fourth (90.48%, 0.630), followed by tracking and feedback systems (80.95%, 0.519). Finally, incorporating diverse physical activities to accommodate varied interests ranked sixth (76.2%, 0.510), stressing inclusivity and sustained engagement.

**Table 12.** Accepted Criteria Position by Ranking for the Element of Physical Activity Habits.

|  | **Condition of Triangular**<br>**Fuzzy Numbers** |  | **Condition of**<br>**Defuzzification**<br>**Process** |  |  |
| --- | --- | --- | --- | --- | --- |
| **Item** | **Threshold**<br>**Value, d** | **Percentage of**<br>**Experts**<br>**Group**<br>**Consensus**<br>**(%)** | **Fuzzy Scores**<br>**(A)** | **Position** | **Expert**<br>**Consensus** |
| How thoroughly<br>are initial and<br>ongoing<br>assessments of<br>physical activity<br>levels integrated<br>into the<br>program? | 0.197 | 95.2% | 0.689 | 3 | Accepted |
| How effectively<br>are physical<br>activity habits<br>integrated with<br>overall health<br>and weight | 0.190 | 85.7% | 0.781 | 1 | Accepted |
| management goals? |  |  |  |  |  |
| Does the program include a wide range of physical activities to cater to different interests and capabilities? | 0.166 | 76.2% | 0.510 | 6 | Accepted |
| Are there effective systems in place for tracking physical activity and providing feedback to participants? | 0.158 | 80.95% | 0.519 | 5 | Accepted |
| How is technology (e.g., apps, wearable devices) used to support and | 0.181 | 85.71% | 0.760 | 2 | Accepted |
| enhance engagement in physical activities. |  |  |  |  |  |
| Are specific metrics used to measure improvements or changes in physical activity habits? | 0.170 | 90.48% | 0.630 | 4 | Accepted |

#### 3.4.4 Element: Dietary Habits

The results from Table 13 prioritize ‘Dietary Habits’ criteria. The highest-ranked criterion, consideration of environmental impacts and sustainability in dietary recommendations, achieved the top fuzzy score (0.768) and expert consensus (90.48%), highlighting alignment with ecological sustainability. Ranked second, aligning dietary habits with individual fitness and weight management goals scored 0.700 with a consensus of 76.2%, emphasizing personalized dietary integration. Thorough assessments of dietary habits during program initiation and progression ranked third (100%, 0.689), underscoring continuous monitoring. Fourth was the consideration of dietary habits’ psychological effects (95.24%, 0.684), followed by ensuring dietary recommendations’ safety based on medical history (81%, 0.648). Lastly, technology use, such as meal-tracking apps, ranked sixth (85.71%, 0.511), enhancing adherence and participant engagement.

**Table 13.** Accepted Criteria Position by Ranking for the Element of Dietary Habits.

|  | **Condition of Triangular**<br>**Fuzzy Numbers** |  | **Condition of**<br>**Defuzzification**<br>**Process** |  |  |
| --- | --- | --- | --- | --- | --- |
| **Item** | **Threshold**<br>**Value, d** | **Percentage**<br>**of Experts**<br>**Group**<br>**Consensus**<br>**(%)** | **Fuzzy Scores**<br>**(A)** | **Position** | **Expert**<br>**Consensus** |
| How thoroughly<br>are initial and<br>ongoing<br>assessments of<br>dietary habits<br>integrated into the<br>program? | 0.195 | 100.0% | 0.689 | 3 | Accepted |
| How effectively<br>are dietary habits<br>aligned with and<br>supportive of<br>individual fitness<br>and weight | 0.154 | 76.2% | 0.700 | 2 | Accepted |
| management goals? |  |  |  |  |  |
| Are the recommended diets safe and suitable for individuals based on their medical history and current health status? | 0.179 | 81.0% | 0.648 | 5 | Accepted |
| How is technology (e.g., meal tracking apps, online nutrition counseling) used to support and enhance adherence to dietary recommendations? | 0.138 | 85.71% | 0.511 | 6 | Accepted |
| Are the effects of dietary habits on psychological well-being considered and addressed? | 0.139 | 95.24% | 0.684 | 4 | Accepted |
| Are environmental impacts and sustainability considered in the dietary recommendations? | 0.156 | 90.48% | 0.768 | 1 | Accepted |

### 3.5 Construct 5: Motivation and Social Support

#### 3.5.1 Element: Motivations for Weight Management

Table 14 highlights the analysis of ‘Motivations for Weight Management’ highlights expert consensus on tailored strategies. Regular feedback and positive reinforcement were ranked highest (fuzzy score 0.700; 76.19% consensus), emphasizing continuous feedback for sustained motivation. Alignment of strategies with individual health goals ranked second (0.698; 100% consensus), reinforcing goal-oriented personalization. Third was tailoring strategies to personal backgrounds (0.665; 76.2%), followed by the clarity of motivational relevance to specific goals (0.640; 85.7%), integration into daily routines (0.592; 85.71%), and adaptability based on changing needs (0.529; 80.95%). These findings support structured, individualized, and adaptable motivational frameworks.

**Table 14.** Accepted Criteria Position by Ranking for the Element of Motivations for Weight Managements.

|  | **Condition of Triangular Fuzzy Numbers** |  | **Condition of Defuzzification Process** |  |  |
| --- | --- | --- | --- | --- | --- |
| **Item** | **Threshold Value, d** | **Percentage of Experts Group Consensus (%)** | **Fuzzy Scores (A)** | **Position** | **Expert Consensus** |
| Are the motivational strategies clear and directly relevant to the individual's weight management goals? | 0.171 | 85.7% | 0.640 | 4 | Accepted |
| How well are motivational strategies aligned with | 0.180 | 100.0% | 0.698 | 2 | Accepted |
| personal health and fitness objectives? |  |  |  |  |  |
| How personalized are the motivational strategies based on individual needs and backgrounds? | 0.174 | 76.2% | 0.665 | 3 | Accepted |
| Are regular feedback and positive reinforcement used as part of the motivational strategies? | 0.154 | 76.19% | 0.700 | 1 | Accepted |
| How well are motivational strategies integrated into participants’ | 0.189 | 85.71% | 0.592 | 5 | Accepted |
| daily routines<br>and lifestyles? |  |  |  |  |  |
| Are<br>motivational<br>strategies<br>flexible and<br>adjustable based<br>on feedback and<br>changing needs? | 0.175 | 80.95% | 0.529 | 6 | Accepted |

### 3.6 Construct 6: Environmental Factors

#### 3.6.1 Element: Environmental Factors

The analysis of grocery shopping habits within “Environmental Factors” highlights their importance in supporting nutritional strategies (Table 15). Personalized shopping strategies aligned with individual dietary preferences ranked highest (fuzzy score 0.768; 90.5% consensus). Feedback mechanisms evaluating shopping habit changes ranked second (0.668; 95.2%), emphasizing regular assessment. Alignment of grocery habits with program nutritional goals ranked third (0.640; 81%), followed by integration with meal planning (0.529; 81%), educational programs promoting healthy choices (0.517; 90.5%), and the influence of shopping behavior on dietary outcomes (0.510; 76.2%). Results indicate personalized, supported, education-oriented grocery strategies enhance dietary success.

**Table 15.** Accepted Criteria Position by Ranking for the Element of Environmental Factors.

|  | **Condition of Triangular**<br>**Fuzzy Numbers** |  | **Condition of**<br>**Defuzzification**<br>**Process** |  |  |
| --- | --- | --- | --- | --- | --- |
| **Item** | **Threshold**<br>**Value, d** | **Percentage of**<br>**Experts**<br>**Group**<br>**Consensus**<br>**(%)** | **Fuzzy Scores**<br>**(A)** | **Position** | **Expert**<br>**Consensus** |
| How well do<br>grocery<br>shopping habits<br>support the<br>nutritional goals<br>set in the<br>program? | 0.180 | 81.0% | 0.640 | 3 | Accepted |
| How<br>significantly do<br>grocery<br>shopping habits<br>influence the<br>dietary choices<br>of participants? | 0.166 | 76.2% | 0.510 | 6 | Accepted |
| Does the program provide adequate education on how to make healthy choices while grocery shopping? | 0.164 | 90.5% | 0.517 | 5 | Accepted |
| Are grocery shopping strategies personalized based on individual dietary needs and preferences? | 0.156 | 90.48% | 0.768 | 1 | Accepted |
| How effectively are grocery shopping habits integrated with meal planning | 0.175 | 80.95% | 0.529 | 4 | Accepted |
| within the<br>program? |  |  |  |  |  |
| Are there<br>feedback<br>mechanisms in<br>place to assess<br>the effectiveness<br>of changes in<br>grocery<br>shopping<br>habits? | 0.128 | 95.24% | 0.668 | 2 | Accepted |

## 4. Discussion

The findings present expert consensus outcomes derived through the FDM. All criteria related to constructs and elements were validated based on agreement among multidisciplinary experts in obesity management, AI, nutrition, psychology, and fitness. This expert consensus informed the development of an integrated framework using the NExGEN-ChatGPT prompt generator for personalized dietary and exercise planning for obese adults. The framework encompasses six primary constructs, subdivided into distinct evaluative elements, and a total of 111 specific criteria, each meeting stringent FDM acceptance standards (>75% expert consensus, threshold value (d) < 0.2, and α-cut > 0.5). These validated criteria collectively form a structured hierarchy model illustrating the integrated AI-based obesity management framework presented in subsequent sections.

The findings from this study provide significant insights into the framework for personalized diet and exercise planning through the NExGEN-ChatGPT system, validated by expert consensus using the FDM. The expert panel identified and prioritized six key constructs, broken down into several elements and criteria, to evaluate the quality of AI-generated plans. These criteria, which met the thresholds of >75% consensus, d < 0.2, and α-cut > 0.5, offer a robust basis for developing and evaluating AI-driven weight management tools. Expert-derived hierarchical models, such as those used in digital-health interventions,^30^ provide a structured approach that helps ensure that the most important quality dimensions (e.g., personalization, safety, and accuracy) are prioritized in system design. Notably, a comparison with real-world outputs reveals a gap between idealized expert models and the actual performance of ChatGPT when generating dietary and exercise plans.^31^ This gap emphasizes the importance of continuous iteration and refinement of the prompt generator to better align AI outputs with expert expectations. Overall, the hierarchical model derived from expert consensus offers a meaningful step towards improving AI integration into personalized obesity care, although further development is needed to ensure consistency with expert recommendations.

The study further highlights the importance of multi-construct frameworks in AI-driven obesity interventions. The top-10 prioritized criteria derived from expert consensus include critical factors such as personalization depth, clarity of content, goal alignment, and safety. These criteria directly correspond to established behavior change theories, such as Goal-Setting Theory and Self-Determination Theory,^32^ which emphasize that goal specificity, user engagement, and empathy significantly influence weight loss success. Research has shown that personalized, clear, and evidence-based recommendations consistently improve the accuracy of chatbot responses, particularly in health domains like nutrition and.^33^ Furthermore, by benchmarking AI-generated diet and exercise plans against clinical guidelines (i.e., from the American College of Sports Medicine and the American Diabetes Association) the study found that while ChatGPT’s advice aligns generally with these standards, certain element like adapting advice to individual medical conditions are occasionally overlooked.^34^ This underscores the necessity of refining the prompt generator to accommodate more specific health profiles and conditions, ensuring that the AI can consistently deliver highly personalized, medically appropriate guidance.

Goal-setting is a central component of successful obesity management, as emphasized by the results of this study, which align with findings from prior research.^35^ Experts highlighted the importance of defining clear, specific, and achievable goals (i.e., based on the SMART criteria) within the NExGEN-ChatGPT framework. The use of specific and measurable targets has been shown to significantly improve engagement and weight-loss success in obese adults.^36^ The study demonstrated that integrating goal-setting with continuous feedback, such as tracking steps via wearables or app-based systems, enhances adherence to diet and exercise plans. These real-time tracking mechanisms are essential in reinforcing goal attainment and adjusting goals dynamically based on user progress. However, as noted in previous studies, there are safety concerns when setting overly ambitious goals for users with specific medical conditions, such as those with joint problems or cardiovascular risks.^37^ By integrating expert feedback into the goal-setting process, the study ensures that targets are both safe and achievable, fostering a sense of accomplishment that promotes long-term adherence.

The findings also support the growing body of research demonstrating the effectiveness of wearables and mobile apps in promoting physical activity among obese adults. Consistent with studies,^38^ the NExGEN-ChatGPT system, when paired with wearable technology, provided a comprehensive tool for tracking activity levels and adjusting exercise recommendations in real-time. The incorporation of technology such as fitness trackers (Fitbit, Garmin) has been linked to improved user engagement, as users are motivated by continuous feedback and real-time reminders to stay active.^39^ Additionally, the study found that personalized, empathetic responses from the chatbot during moments of low motivation or setbacks helped users persist in their exercise routines. This approach aligns with research highlighting the critical role of empathetic, non-judgmental communication in maintaining long-term engagement with weight-loss programs.^33^ In contrast, conflict-prone interactions, where users perceive the AI as overly strict or unempathetic, may lead to disengagement and a decline in physical activity. Therefore, fostering a positive, supportive chatbot-user relationship is essential to sustaining user engagement and ensuring effective behavior change.

The importance of safety and medical sensitivity in AI-generated diet and exercise plans was underscored in the expert consensus. As with earlier research,^40^ the study emphasizes that chatbots must be equipped with robust safeguards to avoid contraindicated recommendations, particularly for users with comorbidities such as diabetes, hypertension, or heart disease. Expert consensus around safety filters and medical history integration was critical in shaping the NExGEN-ChatGPT system, ensuring that recommendations were not only personalized but also medically appropriate. Previous studies have highlighted instances where AI systems, including ChatGPT, delivered potentially harmful advice, especially when users’ medical conditions were not fully considered.^34^ To address this, the study incorporated expert-designed rules for medication-nutrition and medication-exercise interactions, reinforcing the importance of ensuring that AI outputs are aligned with clinical guidelines and best practices. Moreover, privacy and security were prioritized, as safeguarding user health data is essential for fostering trust in AI systems.^36^ These safeguards ensure that the NExGEN-ChatGPT system delivers safe, reliable, and ethical health advice, adhering to both regulatory standards and expert recommendations.

The study also highlights the significance of incorporating lifestyle factors such as sleep, stress, and behavioral habits into personalized obesity management. Research consistently shows that inadequate sleep and chronic stress can undermine weight-loss efforts, with insufficient sleep associated with higher weight regain and poorer adherence to exercise.^35^ The NExGEN-ChatGPT system’s ability to assess and provide tailored advice on these factors is consistent with the holistic approach advocated in recent obesity interventions. By incorporating behavioral health strategies and offering personalized feedback, the chatbot not only addresses physical activity and dietary behaviors but also promotes mental well-being, reducing stress and enhancing motivation. Furthermore, the study aligns with the use of adaptive algorithms in AI systems, where real-time data on sleep, stress, and activity levels enable dynamic adjustments to the user’s plan.^37^ This ability to continuously adapt and provide relevant, personalized support is a key feature of AI-driven obesity interventions and plays a pivotal role in ensuring long-term success.

The findings of this study contribute to the growing body of research on AI-driven obesity management by presenting a robust, expert-validated framework for personalized dietary and exercise plans using the NExGEN-ChatGPT system. This system, grounded in expert consensus, offers a promising tool for enhancing weight management through tailored recommendations, yet its efficacy in real-world settings remains to be fully explored. Future studies should aim to test the NExGEN-ChatGPT system’s effectiveness in diverse populations, including those with varying comorbidities, across different cultural contexts, and with long-term follow-up to assess sustained behavior change and weight loss outcomes. While this study provides valuable insights, its limitations include the reliance on expert consensus without direct user trials, the potential for bias in self-reported data, and the absence of diverse socio-economic and ethnic representation in the expert panel. Further research is needed to validate these findings through randomized controlled trials and to refine the system’s adaptability for broader, more inclusive user groups.

## 5. Conclusion

This study has successfully developed and validated a personalized prompt generation framework using the NExGEN-ChatGPT system, informed by expert consensus through the FDM analysis. The findings highlight the significance of integrating expert-derived quality criteria, such as personalization, safety, and clarity, in the design of AI-driven obesity management tools. The expert panel identified and prioritized critical constructs, which were used to tailor the system’s dietary and exercise recommendations to individual needs. These results demonstrate the potential for AI to support weight management interventions, offering a scalable and accessible alternative to traditional methods. However, while the system’s framework shows promise, the study’s limitations which may include the reliance on expert consensus and the lack of real-world testing obviously underscore the need for further validation. Future research should test the NExGEN-ChatGPT system in diverse populations and real-world settings to assess its long-term efficacy, particularly in terms of sustained weight loss and behavioral change. Additionally, refining the system’s adaptability to individual medical conditions and cultural contexts will be crucial to ensuring its widespread applicability. Overall, this study lays the groundwork for the development of AI-assisted, personalized obesity management tools that can enhance treatment accessibility and effectiveness, providing a valuable tool in the global fight against obesity.

## Acknowledgment

The authors would like to express my heartfelt gratitude to all the experts who contributed their valuable insights to this study. Their expertise in nutrition, exercise, medicine, psychology, and AI has been integral to the development of the NExGEN-ChatGPT framework for personalized weight management. All authors read and approved the final manuscript.

## Declarations

### Author contribution statement

Azwa Suraya Mohd Dan and Adam Linoby: Conceived and designed the experiments; Performed the experiments; Analyzed and interpreted the data; Contributed to data analysis and interpretation of the data; Wrote the paper.

Sazzli Shahlan Kasim, Sufyan Zaki, and Razif Sazali: Conceived and designed the study; Interpreted the data.

Yusandra Md Yusoff, Zulqarnain Nasir, and Amrun Haziq Abidin: Contributed to data analyses.

### Data availability statement

Data will be made available on request.

